# Reconciling motherhood ideology: a classic grounded theory of breastfeeding grief

**DOI:** 10.64898/2026.09.15.26363175

**Authors:** Tumilara Aderibigbe

## Abstract

Breastfeeding grief is a form of complicated and disenfranchised grief which occurs when mothers are unable to achieve their breastfeeding goals; it is characterized by profound sadness and a sense of failure that negatively impact perinatal mental health. The scientific literature consists of very few studies on breastfeeding grief; and a critical gap remains about the primary concern of mothers experiencing breastfeeding grief and how these mothers cope with this concern. This study includes the first substantive grounded theory of breastfeeding grief using a classic grounded theory design. A purposive sample of 46 mothers who are experiencing/have experienced breastfeeding grief completed unstructured virtual individual interviews, and an additional theoretical sample of 19 mothers completed semi-structured individual interviews. Interviews were transcribed verbatim and coded in Quirkos. Data were analyzed using constant comparative analysis, which resulted in the development of a substantive grounded theory. The theory has 3 postulations: 1) the main concern of mothers experiencing breastfeeding is unattained perception of motherhood, 2) the basic psychosocial process used to cope with unattained perception of motherhood is reconciling motherhood ideology, which occurs in 5 stages (responding to disruptions, disavowing reality, grieving, accepting reality, and reframing motherhood ideology), and 3) factors accounting for variation in reconciling motherhood include self-worth, personality, social sentiment/norm, context, and conditions. This theory underscores breastfeeding grief as indeed a perinatal mental health concern, that transcends affective, cognitive, and psychomotor domains. Movement across the stages of reconciling breastfeeding grief is cyclical, not progressive. Therefore, focused psychological interventions to address this concern are urgently warranted.

## Introduction

Mental health issues that mothers experience in the perinatal period including perinatal depression^1^, anxiety^2^, and stress^3^ have been well investigated. However, breastfeeding grief, though commonly experienced by women especially in the early postpartum period, is poorly understood^4^. Breastfeeding grief refers to profound and intense sadness experienced by mothers who did not meet their breastfeeding goals in terms of initiation, duration, exclusivity, or method of milk delivery (direct breastfeeding vs expressed breast milk)^4^. Since introduced in the literature, qualitative descriptions of breastfeeding grief emotions (very sad, depressed, low, guilty, failure etc.) have been reported in the United States^4–7^, United Kingdom^8–10^, and Australia^11,12^. In our previous article^4^, we expanded upon these qualitative descriptions and provided a conceptual formation of breastfeeding grief that included the antecedents, defining attributes, and consequences of breastfeeding grief with which we developed an operational definition. In addition, our finding that breastfeeding grief, being a form of prolonged grief disorder, may be present until 5 years postpartum if unaddressed^4^, corroborates Brown’s finding^8^ that breastfeeding grief may be present until 36 years postpartum. Prolonged and complicated breastfeeding grief results in psychological trauma^8^ and exacerbates postpartum depressive symptoms^7^ (International Classification of Diseases (ICD)-11: 6E20). Whilst findings from these studies improve our understanding of the recently named and understudied concept of breastfeeding grief^5^, a substantive theory of breastfeeding grief is needed to explicate the major psycho-emotional concern with breastfeeding grief and its implications for perinatal mental health.

Further, designs used in previous studies do not directly support the development of a theory about breastfeeding grief, which is needed as a theoretical framework for further studies to fully conceptualize breastfeeding grief towards situating it within the Mental, Behavioral and Neurodevelopmental disorders of the ICD (ICD-11)^13,14^ and the Diagnostic and Statistical Manual of Mental Disorders (DSM). Thus, a significant gap remains regarding a foundational structure for research on breastfeeding grief. Addressing this gap is important because the poor understanding of breastfeeding grief is likely to result in many mothers failing to recognize and seek support for their breastfeeding grief. This is synonymous with disenfranchised grief, that is, grief that is not or cannot be publicly expressed, openly acknowledged, socially supported, or that is misinterpreted or trivialized^15^. Disenfranchised grief can exacerbate, disrupt, or perplex the process of emotional recovery^16^. It can also cause individuals to become estranged from outside resources, strain relationships, and result in depression^17,18^. Often times, disenfranchised grief transcends into complicated/prolonged grief disorder^19^ (grief that persists for more than 12 months^20^).

Unlike most types of grief, breastfeeding grief is unique in that it is not associated with death of a human rather it is the ‘death/loss of breastfeeding experience’. Hence, existing theories of grief are not suitable for understanding breastfeeding grief. A specific theory of breastfeeding grief is thus required to elucidate breastfeeding grief using a data-driven approach to generate a robust and nuanced theory. Findings from this study will contribute to the body of knowledge on perinatal mental health and promote understanding of the trajectory of breastfeeding grief and factors influencing it. Findings from this study will also serve as preliminary data for future studies on breastfeeding grief to promote perinatal mental well-being.

The aim of this study was to generate a substantive theory of breastfeeding grief. In accordance with Glaser’s classic Grounded Theory’s ‘no preconception dictum’^21,22^ we set to facilitate a *natural emergence* of a substantive theory, or explanatory model, that is specific to our study population. A literature review of qualitative studies reported in this study was conducted after our substantive theory was generated. Our broad/open research questions^23,24^were:

1) What is the main concern of mothers experiencing breastfeeding grief?
2) What is the basic social process that mothers use to process the main concern?
3) What factors account for variation in processing the main concern?

## Methods

### Philosophical Underpinning

This study has its philosophical underpinning in symbolic interactionism. Originating from George Herbert Mead^25^, symbolic interactionism describes the process through which humans create and maintain social structures using meaningful symbolic communication^26^. We used symbolic interaction to understand the construed meaning of the main concern experienced by mothers and how this concern is influenced by societal interaction.

### Design

Qualitative Classic Grounded Theory was used^27^ in line with the aim of the study to develop a substantive grounded theory of breastfeeding grief. We drew from Glaser’s classic methods over the other two variants of grounded theory – Straussian^28^ and Constructivist^29^ – to generate a theory free from researchers’ preconceptions^23^. Straussian grounded theory was developed by Strauss and Corbin and Constructivist grounded theory was developed by Charmaz. Both methods emphasize the researcher’s active role in data interpretation and theory generation. Additionally, we used classic grounded theory following Stern’s^30^ recommendation that classic grounded theory is suitable to examine a complex health concern when: a) little is known about a topic, b) there are no pre-existing theories or existing theories do not adequately describe the topic. Stern further added that in this case, grounded theory affords the researcher to build an explanatory theory directly from the data collected^30^. Applied to the present study, breastfeeding grief is understudied and existing grief theories are not suitable to examine it because it does not involve the death of a ‘human.’

The non-linear steps of classic grounded theory were followed, including simultaneous data collection and analysis, substantive coding (open and selective coding), constant comparison, theoretical sampling, theoretical coding, memoing (conceptual and theoretical), sorting^31^. These steps guided the emergence of participants’ main concern and the core category of this substantive theory of breastfeeding grief. The study was conducted and reported according to ‘Developing a Guideline for Reporting and Evaluating Grounded Theory Research Studies’ (GUREGT) (Supplement 1)^32^.

### Recruitment

Passive recruitment approach was employed using printed and virtual research flyers^33^ from February 2026 to September 2026. Printed flyers were distributed at the University of Massachusetts (UMass) Chan OBGYN unit, UMass Boston campus and greater Boston area. Following ethical guidelines for social media recruitment^34^, virtual research flyers were posted on Facebook (Facebook groups for moms and Facebook ads targeting moms) and Instagram. Participants were also recruited through ResearchMatch^35^ and community and professional organizations including Postpartum Support International (PSI)^36^, Black Mamas Matter Alliance (BMMA)^37^, and the Institute for New England Native American Studies (INENAS)^38^ at UMass Boston.

### Participants

Data from 65 participants in the United States were analyzed. Following Glaser’s guideline that a classic grounded theory should be developed with both purposive and theoretical sample^39,40^, a purposive sample of 46 mothers who met the following inclusion criteria were included. Biological women who: (a) are 18 years or older (eligible participants must be 19 years or older if living in Alabama and Nebraska, and 21 years if living in Mississippi); (b) have access to Zoom® and can complete an online survey; (c) can speak and read English; (d) have a biological child who is alive; and (e) is experiencing breastfeeding grief (deep sadness/grief due to unmet breastfeeding goals for the child). Exclusion criteria will be women: (a) < 18 years of age; (b) with a diagnosis of serious mental health disorder e.g., schizophrenia, bipolar disorder; and (c) have experienced perinatal, neonatal, or infant loss.

An additional theoretical sample of 19 mothers who met the same inclusion criteria above (with the exception of having a biological child under 1 year) were included to shape the theory. Details about revisions to the screening questions are included in Supplement 2.

### Setting

Data were collected virtually from mothers residing in the United States, using Zoom® for interviews and Research Electronic Data Capture (REDCap) screening survey for surveys^41^.

### Data collection procedure

Recruitment flier contained the research team’s email address and a QR code leading to the REDCap screening survey; 168 potential participants completed the screening survey for the purposive sample, out of which 100 were identified as eligible. Eligible participants were sent an email containing details about the study, informed consent form, consent form for recording and transcribing, and a request to indicate interest in participating in the study after reviewing the informed consent form. Participants who indicated interest were contacted via phone call to verify answers to screening questions and schedule the online interview. Of the 100 eligible participants, only 46 responded to the follow up phone call and completed the interview. For the theoretical sample, 125 potential participants completed screening of which only 21 responded to the follow up phone call and completed the interview. Hence, a total of 65 participants were enrolled for the grounded theory. Individual in-depth interviews were conducted via Zoom® by the author following guidelines for conducting interviews via video conferencing^42^. Individual interviews were chosen over focus groups because a) the primary data collection for classic grounded theory is interviews^43^, b) breastfeeding grief is a sensitive topic^44^, c) interviews afford the researcher to process gestures, pauses, and deep emotional context that may be missed in a focus group^45^. Before each interview commenced, the interviewer reviewed the informed consent form, answered questions and confirmed verbally that participants still wished to participate. The interviewer also initiated a researcher-participant relationship to build trust and rapport.

### Data collection and analysis (initial purposive sample)

Data collection and analysis in classic grounded theory occur simultaneously^24,27^ through the process of constant comparative analysis^46,47^. Unstructured individual interviews were conducted with the purposive sample by asking a *grand tour* question – *Can you please share your breastfeeding experience with me?* followed by probing or clarifying questions (e.g., you mentioned that you fed your child with other foods apart from breast milk, can you please share what these were?). Unstructured interviews were audio recorded and lasted an average of 22 (range: 9-41) minutes. The goal of this study was to generate a theory transferable to mothers across the United States, hence, we enrolled participants across sociodemographic groups and until the core category emerged^27,31^. After each interview, transcripts were downloaded from Zoom and reviewed by the author and research assistants.

Glaser’s 4 steps of constant comparative analysis were followed, and the author wrote case-based memos reflecting on what transpired during the interview and throughout the study^27,46^. 1) *Comparing incidents applicable to each category*: Reviewed transcripts were uploaded in Quirkos (web version) for coding^31^ followed by line-by-line open coding by TA. Open coding involves a line-by-line conceptualization of descriptive incidents within the raw data and comparing them to each other to identify substantive codes and the shared concerns of mothers and the core category^48^. During open coding, incidents were initially coded into 37 substantive categories^46,48^. (Table 1). Concepts with low number of incidents were merged (in our case, less than four). Hence, ‘motivating’ was merged with ‘affirming,’ ‘observing’ was merged with ‘knowing,’ ‘yearning’ was merged with ’acknowledging,’ and ‘self-judging’ was merged with ‘reacting.’ The revised number of substantive codes was 33. Incidents were compared to previous incidents coded in the same category to examine similarities and differences^39^ and this process resulted in the identification of theoretical properties of categories^46^. TA created memos in a Word document about possible properties of each category^46^. 2) *Integrating categories and their properties*: Incidents were compared with the properties of each category, revealing the relationships among properties of a category and relationships among categories^46^. Memos about the connections among categories and connections among properties of categories were created in Word^48^. As categories and their properties were integrated, a core category – grieving (conceptualized from reacting) – began to emerge, which seemed to account for most of the variation in the pattern of participants’ behaviors^48,49^. 3) *Delimiting the theory:* After identifying the core category, the theory was narrowed to 7 conceptual categories (anticipating, struggling, acknowledging, reacting, comparing, internalizing, accepting) that have explanatory power and are relevant to generate the theory while remaining theoretically sensitive. This facilitated generating a parsimonious theory that accounts for the maximum variation in behavior using the minimum number of concepts. The theory was delimited to focus on how participants continually process/resolve their main concern and selective coding of incidents in these 7 categories was conducted while remaining theoretically sensitive^49^. Selective coding saturated the core category and related categories; hence, the first phase of data collection was completed.

**Table 1.** List of the original 37 substantive/open codes.

|  |  |  |
| --- | --- | --- |
| Reacting | Self-judging | Reflecting |
| Attributing | Opining | Re-strategizing |
| Affirming | Explaining | Advocating |
| Acknowledging | Timing | Justifying |
| Characterizing | Blaming | Valuing |
| Comparing | Questioning | Wishing |
| Recounting | Connecting | Understanding |
| Knowing | Appreciating | Complying |
| Rationalizing | Attempting | Accepting |
| Struggling | Predicting | Motivating |
| Anticipating | Coping | Observing |
| Experiencing | Internalizing | Yearning |
| Perceiving motherhood |  |  |

### Data collection and analysis (theoretical sample)

3) *Delimiting the theory [continued]:* Data from a theoretical sample^40,49^ is required to further delimit the theory therefore, semi-structured individual interviews were conducted with participants who have children under 1 year as incidents from the purposive sample suggest that breastfeeding grief is most intense in the first year postpartum. Interviews were conducted by the first author using an interview guide containing questions to tease out concepts in the emerging theory (Supplement 3). Interviews were audio recorded and lasted approximately 21 (range: 9-34) minutes. Transcripts were uploaded in Quirkos and theoretical coding was employed to understand the theoretical connections between the core category, its properties, and relevant categories^40,49^.TA read Glaser’s *families of theoretical codes* to become more theoretically sensitive to the data^50,51^ and be empowered to “generate theory and keep its conceptual level”^52^. Hernandez (2009)^50^ noted that several theoretical codes may emerge in a grounded theory study however, the most relevant theoretical code will be the one that captures the relationship with the core category and related categories. Indeed, five theoretical codes emerged from our data: from the process family^49^ (stages), from the basics family (basic social psychological process), from the 6 C’s^49^ (causes, consequences), and from the cultural family (social sentiment). Further examination of these theoretical codes revealed that ‘*basic social psychological process*’ best captures the relationship between the core category and related categories. Theoretical saturation was reached after interviewing the 65^th^ participant^40,53^. Therefore, an explanatory model of the relationships between the main concern and its properties, and the main concern and other related categories (supported with memoing), led to substantive theory generation as illustrated in figure 1^48^. 4) *Writing the theory:* Glaser’s guidelines on theoretical writing were followed to ensure that the theory is not descriptive, but rather conceptual^54,55^. Memos on grieving and related categories were sorted for an outline^48,54^. Categories were reported as major themes with memo discussions constituting their contents. Relevant theoretical codes (self-worth, social sentiment/norm, context, and conditions) which are factors accounting for variation in the processing the main concern were reported in the results.

**Figure 1.**
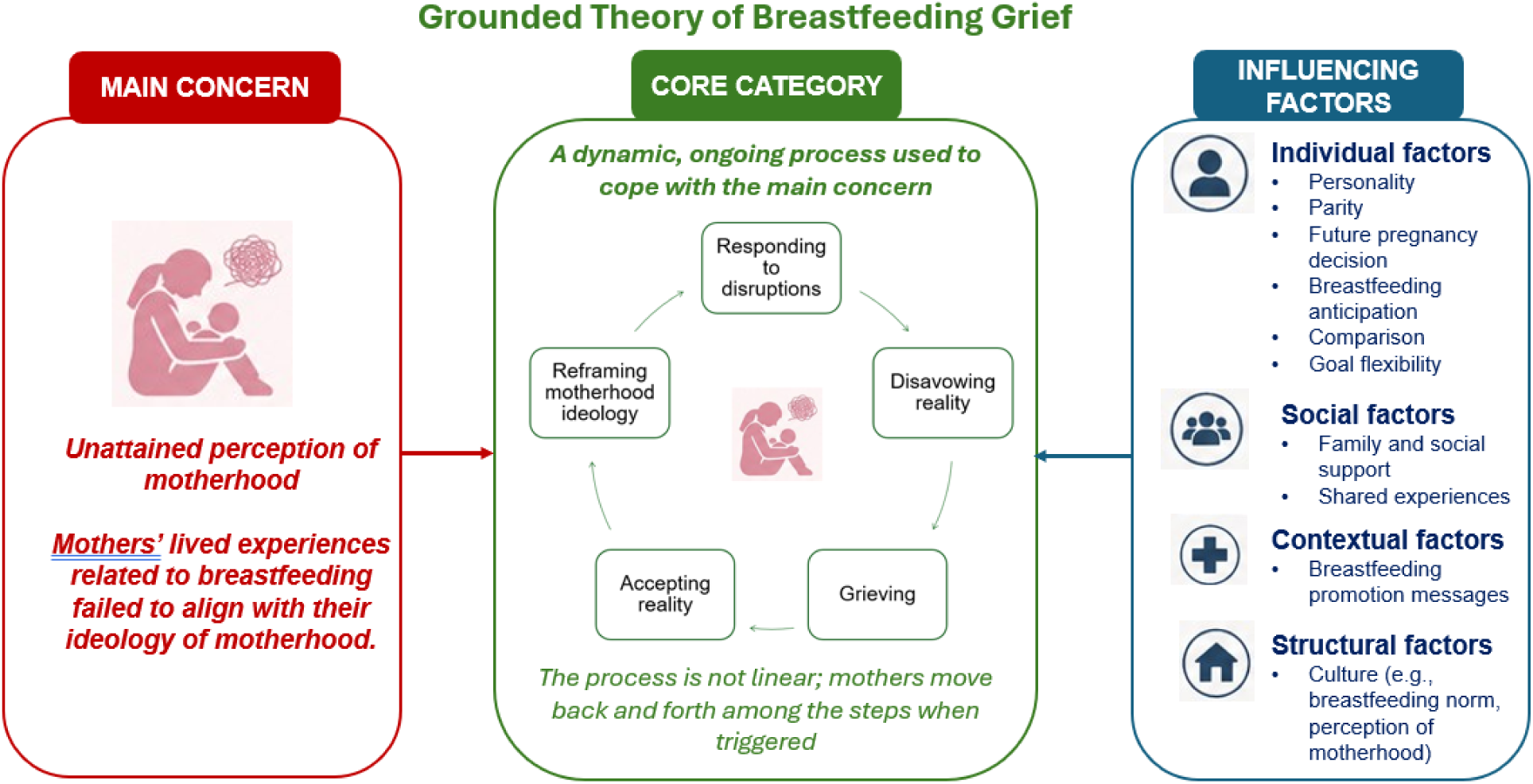
Substantive grounded theory of breastfeeding grief, highlighting the main concern, core category, and factors that account for variation in processing the main concern.

#### Rigor

We followed Glaser’s recommendations for conducting and writing a classic grounded theory^27,54^. Memos were created throughout the data analysis process to guide the theory generation and writing^56^. Further, we followed Berthelsen and colleagues’ guidelines for reporting classic grounded theory studies^32^. We evaluated our substantive theory based on Glaser’s eight criteria: fit, understandability, relevance, grab, general, work, control, and modifiability^27,49,51^. The theory closely *fits* the data and the substantive area of breastfeeding grief, as concepts were derived directly from the data and not preconceived. We presented the theory in simple terms that both professionals and laypersons involved with breastfeeding can *understand*. Our theory is *relevant* because it explains the main concern of mothers who did not meet their breastfeeding goals. The interesting relationships between concepts in the theory facilitates the theory’s capability to *grab* the attention of people interested in breastfeeding. The wide scope of the main concern ‘unattained perception of motherhood’ makes our theory *general* enough to be applied to any concern in this substantive area^57^. We defined unmet breastfeeding goals in terms of initiation, duration, exclusivity, or method of milk delivery, this makes our theory suitable to any of these four goals. In addition, our substantive theory *works* through the explanation of what happened, the interpretation of what is happening and the prediction of what is happening when mothers do not meet their breastfeeding goals. This allows us to have *control* over the situation being examined. Finally, our emerging theory can be modified as new data on mothers’ experiences of not meeting their breastfeeding goals become available^49^.

### Ethical considerations

The study was approved by the University of Massachusetts Boston Institutional Review Board (IRB#: 4328). Confidentiality and data security were ensured by storing data in the University’s secure cloud storage. Interviews were conducted using HIPAA-compliant Zoom.

## Results

A substantive grounded theory of breastfeeding grief was developed. The theory has 3 postulations: 1) the main concern of mothers experiencing breastfeeding grief is *unattained perception of motherhood*, 2) basic social psychological process to navigate this concern – reconciling motherhood ideology – which occurs in five stages: a) responding to disruptions (when mothers experience emotional and psychological shock owing to unexpected challenges that are highly likely alter their breastfeeding goals); disavowing reality (when mothers are faced with the glaring fact that their breastfeeding goals need to be altered urgently but simultaneously acts as if it is untrue, to reconcile conflicting emotions); grieving (when mothers express how they truly feel about not meeting their breastfeeding goals); accepting reality (when mothers accept that they did not meet their breastfeeding goals); and reframing motherhood ideology (when mothers change their perceptions about their main concern by reconceptualizing it to have a balanced, empowering interpretation), and 3) factors that account for variation in reconciling motherhood ideology include self-worth, personality, social sentiment/norm, context, and conditions.

### Main concern

Though mothers have shared concerns including the loss of the breastfeeding experience they had imagined, worry about child’s health, and concerns about experiencing breastfeeding difficulties for future pregnancies, their main concern is *unattained perception of motherhood*. This concern was not preconceived, rather it was identified directly from the raw data through open coding. In other words, they were concerned about qualifying as a ‘good mother’ after not meeting their breastfeeding goals. A mother affirmed, “I felt like I wasn’t a good enough mother.” And another remarked, “I felt like I’ve lost that version of early motherhood that I had dreamed about and tried as much as possible to, you know, um to make it a reality.” Historically, the word ‘mother’ is perceived in relation to nurturing her young. Hence, women who hold this title (as a result of having biological children) are often expected to love and nurture their children. For mothers with infants, this nurturing responsibility is breastfeeding. Hence, breastfeeding is largely perceived as a yardstick for motherhood. Therefore, when mothers’ lived experiences related to breastfeeding fail to align with their ideology of motherhood, it becomes concerning for them.

### Core category

Going forward in the process of open coding, the core category of *reconciling motherhood ideology* emerged, which has the most explanatory power in the theory and explains the process that mothers go through to continually process or resolve their main concern. Reiterating that mothers indeed process the main concern, a mother affirmed, “It’s still an ongoing process, or, like, I’m still processing the situation.” Constant comparative analysis suggests that *reconciling motherhood ideology* is the cognitive process through which mothers gradually acknowledge the psychological discomfort of not meeting their breastfeeding goals and redefine their ideology of motherhood.

#### STAGE 1: RESPONDING TO DISRUPTIONS

The first stage of the theory of breastfeeding grief is *responding to disruptions*. This stage is characterized by emotional shock and psychological shock, which occur following a negative trigger (in this case, unexpected challenges related to breastfeeding). The mind remains in a passive state, especially when it is filled with positive predictions about a situation (in this case, breastfeeding), until a negative stimulus is experienced, which propels it to an active state. The immediate response to this active state is emotional and psychological shock, manifested inwardly or outwardly as anger (intrapersonal or interpersonal), panic, embarrassment, awkwardness. Affirming this stage, a mother stated, “I was in this state of you know, um shock.” Another mentioned, “It was just like, such a sudden event. It was so quick and so intense. “ As mothers progress through stage 1, they experience the properties of responding to disruptions as they pivot: *expressing alarm, reflecting on predictions, and acknowledging dissonance*.

### Expressing alarm

The sudden violation of expectancy triggers the first survival response of alarm which signals loss of control. A mother mentioned, “It just felt abrupt and um out of my control.” Mothers express shock at the disruptions, particularly because this is in opposition to their existing breastfeeding expectations. This state of destabilization manifests as internal and/or external chaos (dimensions) that challenges maternal self-efficacy, as expressed in the statement “When I just couldn’t continue, you know, it felt like a hit to my confidence as a new mom.” Internal chaos destabilizes the mind with cascading influx of automatic cognitive processes and thoughts whereas external chaos is characterized by behavioral volatility. Destabilized mothers move back and forth between these intertwined forms of chaos as they express alarm and attempt to process the disruptions.

### Reflecting on predictions

Breastfeeding expectations come to mind again in an attempt to make sense of the disruptions, as illustrated in this reflection: “I pictured it being very natural, you know, most magical moment, like my baby would latch right away and would just know what to do.“ Reflecting on predictions is also a cognitive reality-checking mechanism of searching the memory to verify that they have not been previously warned of the disruptions as they were anticipating a positive breastfeeding experience. This provides a logical explanation for experiencing shock.

### Acknowledging dissonance

Still responding to disruptions, cognitive dissonance is experienced, which involves psychological discomfort due to unmet predictions, and is characterized by acknowledging discrepancies between predictions and actual experiences. One mother stated, “When he did eventually latch, it was painful very painful. Like it was like there’s a blade trying to someone using a blade trying to you know, kind of scrape off my skin. That was how painful it was. I wasn’t, you know, expecting that level of discomfort.” Acknowledging dissonance begins the realization that a goal may be unmet and this can be scary for mothers. In an attempt to wade off this realization, mothers move to the next stage of disavowing reality.

#### STAGE 2: DISAVOWING REALITY

Disavowing reality functions as a psychological defense mechanism against cognitive dissonance. This unconscious process is aimed at resolving emotional conflict hence, it highlights the core category – grieving – as indeed the strategy that mothers use to process/resolve their main concern. It is a state characterized by rigidness of the mind about a painful reality. A mother expressed, “I was in denial. I was in denial that no, like this can’t be happening to me.” As they move through stage 2, they attempt to disavow the reality of their unmet expectations through *rationalizing* and *questioning*, which are conceptualized as subcategories.

### Rationalizing

Recounting actions taken to achieve their breastfeeding goals is one of the ways that mothers provide logical explanations for why they shouldn’t have experienced disruptions that resulted in unmet goals. A dimension of rationalizing is audience (intrapersonal vs interpersonal). Mothers self-rationalize to protect their ego and reduce cognitive dissonance, as they convince themselves that not meeting their breastfeeding goals isn’t their fault (or at least entirely) because they took preventive measures. One mother remarked, “I wasn’t expecting to go through this challenge. So, I was in denial. I was in denial that no, like this can’t be happening to me. Like I prepared myself. I went to the workshops. I, I have my tools that I need. I’m ready. I’m prepared.” Like intrapersonal rationalization, interpersonal rationalization is also an attempt to protect ego, however, it is focused on preventing external blame related to an unmet breastfeeding goal. “I would always breastfeed in front of my son’s father to almost like prove and show like, look, I am trying. Like, you don’t think I want to be a good mom? Like, I want to be able to feed our child. I want to be able to feed our son, but it’s like, okay, what am I supposed to do? Like, there’s no milk coming out. Like, I took this medicine. Okay, they’re recommending this herb and they’re recommending this and that.”, a mother stated.

### Deflecting

Further disavowing the painful reality of their unmet breastfeeding goals, mothers ask themselves several questions, to evade the reality. A mother asked, “I had to start having a flashback and rethinking what did I eat that make me that’s making me this way? What did I eat? What? Is it my cravings? Because I have really weird cravings in my pregnancy” The questions may be directed at self or others (dimension) as mothers continue to shift their attention away from the painful reality. Questions directed at self, take the form of introspective questioning to evaluate individual actions that may have resulted in an unmet breastfeeding goal. Extrospective questioning directed at others is also a means to avoid the painful reality of unmet breastfeeding goal, as illustrated in a mother’s statement, “What’s wrong with him [my baby]?”

#### STAGE 3: GRIEVING

Ruminating looping thoughts about the dissonance reach a peak, resulting in emotional meltdown. This behavioral response functions as an active multidimensional process where emotional balance is disrupted, due to the destabilization created by an unmet goal. “My baby is only five months old and I’m still kind of grieving it.” a mother said. Grieving reinforces the fact that the premature loss of anticipated experience is deeply personal and very valuable, as a mother illustrated, “It’s like a quiet loss that doesn’t get, you know, talked about much because my baby is alive and healthy” Grieving, a highly individualized and non-linear process, also begins the adaptive process of processing the main concern towards moving to acceptance and healing. A mother opined, “I feel that to heal of something that came as a shock to you as a person it’s very hard,” nonetheless, healing can be achieved through the defining properties of this active stage include *emotional meltdown, isolating, questioning, blaming, catastrophizing, and wishing*.

### Emotional meltdown

The painful reality of an unmet goal disrupts emotional composure leading to torrents of emotions including sadness, anger, moodiness, depression, guilt, failure, disappointment, frustration, defeat, and irritability. These emotions can be adaptive or maladaptive (dimension) and may manifest as internal distress only or involve outward expression (dimension). Another dimension is the intensity (mild to severe) and duration (transient vs prolonged). Adaptive emotions are natural and flexible (often transient) and are aimed at processing a painful experience towards achieving stabilization. Maladaptive emotions on the other hand are prolonged (lasting more than 1 year) and negatively impact functioning. Narrating her experience, a mother mentioned “I couldn’t give my 100% to anything. I couldn’t give my 100% to work. I couldn’t give my 100% to my child or to my husband.” Mild emotions tend to be transient and adaptive whereas severe emotions are prolonged and often maladaptive. Emotional meltdown, manifested as internal distress only, is both a function of personality trait referred to as ‘overcontrol’, and stigma related to unmet expectations. Overcontrol is characterized by excessive emotional suppression and self-regulation. Outward expressions of emotional meltdown are catalyst for healing; this may be personal or interpersonal.

### Isolating

Isolating provides the opportunity to process the painful reality in solitude; it also acts as a psychological defense mechanism against shame and emotional meltdown. Withdrawal protects the ego and acts as a barrier to unwanted social intrusions that may reinforce emotional meltdown. Isolating may be short-term or long-term (dimension) and digital or non-digital (dimension). Short-term withdrawal signifies quicker movement to reframing while long-term withdrawal indicates a slower route to reframing. Stating the duration of her isolation, a mother remarked, “For about two weeks I didn’t really speak to anybody.”

### Questioning

The dissonance between expectations and reality results in frantic efforts to understand the pathway to the painful reality, hence, questioning is employed for this purpose. The intent of questioning may be positive/adaptive or negative/maladaptive (dimension). When the goal of questioning is to understand the pathway to prevent future recurrence, this is helpful towards speedy progression to reframing, as indicated in a mother’s comment, “Should I have pumped more, you know, drank more water? (sigh) probably seen another lactation consultant?” Conversely, maladaptive questions are biased, undermine problem-solving, and are aimed at blaming, as reflected in a mother’s comment, “I look at my chest and I’m like, why do you guys decide to disappoint me?”

### Blaming

One of the outcomes of maladaptive questioning is blaming, which is directed at self or others (dimension). Because breastfeeding is perceived as an essential mothering role, blaming is often self-directed and highly critical, involving statements like, “I already failed my first lesson as a mom.” as a mother lamented. Blaming delays progression to reframing and healing but brings temporarily relief when externally directed, as guilt is projected onto someone else.

### Catastrophizing

Catastrophizing magnifies the painful reality; hence, it is a maladaptive coping mechanism with negative predictions for the future. Another mother’s self-critique was, “I failed at something that was supposed to be fundamental, you know, that I had to experience, that is part of life, life itself, the core of life.” Labeling is one of the features of catastrophizing, as indicated in a mother’s comment, “I felt basically like a failure.” Catastrophizing negatively impacts self-esteem and opens up the mind to accept future recurrence of negative events.

### Wishing

As a property of grieving, wishing is a medium for reflecting on strategies that could have prevented dissonance between expectations and realities and/or strategies to navigate emotional meltdown. This is reflected in a mother’s remark, “I wish I could have had more prep about breastfeeding” The intensity of reflection and accompanying thoughts can either provide temporary relief or exacerbate emotional meltdown. Wishing may be personal or interpersonal (dimension), and it slowly moves mothers towards accepting the painful reality.

#### STAGE 4: ACCEPTING REALITY

This transitional phase marks the beginning of healing; it is an active stage where emotional meltdown softens, and the permanent reality of the loss is acknowledged. Affirming the permanence of the loss, a mother stated, “That was a lost opportunity that I can never get back again.” Acceptance is the catalyst that transforms the grief from a state of disavowal into the state of emotional healing and cognitive clarity, however, it does not lessen the significance of the loss as illustrated in a mother’s remark, “healing doesn’t mean, you know, pretending that it didn’t matter.” Movement through this phase involves experiencing its properties including *conceding*, *resolution*, and *integrating*, which are conceptualized as subcategories.

### Conceding

Grieving encompasses emotional release and curtails maladaptive rumination; this in turn increases the cognitive clarity needed for concession. Conceding may be direct or indirect (dimension), nonetheless, its goal is to facilitate acceptance of the painful reality towards stability and rebuilding. Ego defenses drop during conceding as mothers relinquish the illusion of control, reflected in a mother’s remark, “I see healing as, you know, allowing myself to acknowledge that this was important to me and that it is okay to grieve it.” Conceding also provides a level of emotional and mental relief from the stress of disavowing reality and initiates the active pathway to growth and genuine healing.

### Resolution

Conceding the painful reality brings renewed hope through cognitive processing of the situation, and aids resolution. Finding an emotional closure is a defining feature of resolution that advances the healing journey, and it may originate from self or others (dimension). A mother confirmed her personal resolution saying, “All we just need personally is a closure, like something that we can part with just to stop feeling the way I am feeling.” Another mother stated that the closure is dependent on her child, “My baby doesn’t have any illness at all. He’s very, very healthy. So, I think I’m very, very fine. I’m okay. I’m no longer unhappy.”

### Integrating

Finding a closure provides the mental capacity to integrate the loss into one’s identity, referred to as ‘identity adjustment.’ This also reduces the shame associated with the painful reality and increases self-esteem that was otherwise impacted during *catastrophizing*. Integrating is a gradual psychological evolution where the loss is repositioned in the mind as part of one’s identity rather than an open, prolonged traumatic experience. The painful reality becomes manageable as this is achieved and enables a quick progression into the final stage of healing – reframing.

#### STAGE 5: REFRAMING MOTHERHOOD IDEOLOGY

Though acceptance brings renewed hope and is characterized by reduced intensity of emotional meltdown, the process of healing is not complete until the main concern is addressed. This highlights the importance of the final stage where motherhood ideology is reframed and resilience is improved. Altering initial perception about motherhood fractures rigid perspectives underlying the main concern, and this may be done individually or collaboratively (dimension). It is marked by a shift from *blaming* to self-compassion, and this is succinctly captured in the comment, “I feel like I’m slowly moving from guilt and self-blame, you know, towards acceptance and compassion for myself.” Reframing is an internal process that may be limited to internal processing only or externally expressed (dimension). A mother’s active and independent role in reframing is exemplified in her saying, “I fed my baby in the way that kept him growing and healthy, you know, that’s still being a good mom, that is still love.” Reframing is a cognitive process defined by properties including *reorienting, meaning making*, and *identity reconstruction*.

### Reorienting

Reorienting involves real-world adjustments to incorporate the new mindset acquired through reframing. It involves breaking down reframing, which is ‘abstract,’ into concrete actions and the practical execution of those actions in daily routines. This active process is individualized and sustains healing. “Instead of saying that I failed, you know, I’m trying to reassure myself that I didn’t fail. So, I’m just trying to say I adapted to um new beginnings,” said a mother about her reorienting strategy. Reorienting is basically learning to live in a ‘changed’ world.

### Meaning making

Meaning making is an active endeavor to understand the painful experience in its totality, and how this applies to one’s existence. It involves searching for new meanings within the accepted reality to facilitate post-traumatic growth and improve emotional regulation. Reflections and critical analysis conducted during meaning making are aimed at generating a cohesive story about the experience to find purpose, like a mother described, “I understand that sometimes things don’t go according to plan, and I know that feeding my baby and keeping my baby healthy was most important thing. That is my priority right now.”

### Identity reconstruction

New meanings generated during meaning making are crucial for identity reconstruction. Negative perspectives about self-identity are modified to enhance growth and increase self-worth. Identity reconstruction involves rebuilding self-image and also incorporates new tasks or roles to achieve that purpose. A mother captured this in her saying, “If your goal was to breastfeed and you tried breastfeeding, you’ve already been a great mom for your child by making sure that that’s your priority, at least trying it.”

### Factors that account for variation in processing the main concern

The core category (reconciling motherhood ideology) is the central strategy through which mothers resolved their main concern, this process is not utilized in the same way due to individual differences. These include self-worth, personality, social sentiment/norm, context, and conditions. Mothers with low self-esteem, type A personality, cultural norm of breastfeeding, and those with higher exposure to successful breastfeeding experiences or contents, reported these as triggers/influencers for the process used to navigate the main concern.

## Discussion

The Glaserian classic grounded theory of breastfeeding grief that emerged from this study, *Reconciling Motherhood Ideology*, conceptualizes breastfeeding grief experiences of mothers identifying the main concern as unattained perception of motherhood. Mothers navigate this main concern through a process referred to as reconciling motherhood ideology, which consists of five interconnected stages: *Responding to Disruptions, Disavowing Reality, Grieving, Accepting Reality, and Reframing Motherhood Ideology*. These evolving stages comprise emotional, behavioral, and cognitive adjustments that move from emotional destabilization to balance and reorientation. The stages of grieving revealed that some mothers experience prolonged maladaptive emotions and coping mechanisms that render them fixated as they internalize their grief. This positions breastfeeding grief as a form of prolonged grief disorder according to the DSM (DSM-5-TR:F43.81) and ICD (ICD-11:6B42). Further, Carlson and Dalenberg (2000)^58^ identified three defining features of a traumatic event including loss of control over the situation, perceiving that the event is a highly negative experience, and the suddenness of the experience. These features are present in our theory hence breastfeeding grief qualifies as a traumatic event.

In this theory, grieving is one of the stages involved in reconciling motherhood, whereas *Kübler-Ross model* conceptualized it as a central phenomenon in loss and explained it in stages^59^. The properties of the stages of reconciling motherhood in the present theory are similar to those reported in previous grief-related theories including *Kübler-Ross model*^59^*, Theory of care realignment^60^*, *Dual Process Model of Grief^61^*, and *Meaning reconstruction theory*^62^. For example, the stage of responding to disruptions describes the initial emotional destabilization that occurs subsequent to disruptions to predictions whereas disavowing reality is characterized by illusion. Both stages are merged as ‘denial’ in *Kübler-Ross model*. The third stage of the theory of breastfeeding grief is grieving, characterized by emotional and behavioral collapse; these are reflected in the ‘anger,’ ‘bargaining’ and ‘depression’ stages of *Kübler-Ross model* and as properties of the phase ‘experiencing loss,’ in the *Theory of care realignment*. Acceptance is one of the stages in *Kübler-Ross model;* it is also included in the theory of breastfeeding grief and conceptualized as ‘accepting reality.’ The stage of reframing motherhood ideology is similar to the phases of ‘reorienting’ and ‘rebuilding’ in the *Theory of care realignment,* where individuals who are experiencing grief due to loss of life, embark on the journey of healing, self-preservation, and reshaping routines and personal identities*^60^*.

The theory of breastfeeding grief’s unique contribution to the literature on grief is the postulation that the experience of grief is not limited to loss of human life, rather, grief may be present in any experience involving a loss, especially a premature loss. Further, the theory of breastfeeding grief addresses a distinct main concern – unattained perception of motherhood which is often internalized. This theory thus provides an opportunity to promulgate this perinatal mental health concern; hence, the properties of reframing are focused on this concern as opposed to those in existing grief theories which are focused on managing life after loss of a loved one. Considering individual variability in adaptation, the theory of breastfeeding grief is not linear, and individuals may move among the stages, experience multiple stages concurrently, or not experience some stages. This is congruent with as in *Kübler-Ross model* and the *Theory of care alignment*.

Overall, the theory of breastfeeding grief identifies that grief is not solely an experience for individuals who have experienced loss of human life, rather, individuals who have experienced any form of loss. It recognizes motherhood ideology as central to breastfeeding grief, and highlights that a core element for healing is reframing as in *Meaning reconstruction theory*.

## Strengths and Limitations

There are many strengths to this study. First, a novel substantive grounded theory of breastfeeding grief was generated, which provides a theoretical framework for future research on breastfeeding grief. Second, the theory is general and may be applied in different circumstances of unmet breastfeeding goal (i.e., initiation, duration, exclusivity, and mode of milk delivery). Participants from several sociodemographic backgrounds were included, and this increases the theory’s transferability. Nonetheless, there are some limitations. The purposive sample included participants with past experience of breastfeeding grief, which may introduce recall bias. However, compared to theoretical sample with recent experience, the purposive sample provided more nuanced insight into their breastfeeding grief experiences. This may be because the brain processes hindsight with complete context. Average interview duration for theoretical sample was shorter compared to purposive; this may be due to the demands of the postpartum period, especially sleep deprivation, energy demands of breastfeeding, caring for infant etc.

## Implications

This theory has implications for clinical practice and research. The theory describes ‘what is going on’ with mothers who are experiencing breastfeeding grief, towards conceptualizing breastfeeding grief and differentiating it from other perinatal mental health issues such as postpartum depression. It is possible that mothers experiencing breastfeeding grief may have been misdiagnosed as having postpartum depression or other mood disorders, since very little is known about breastfeeding grief. In addition, these mothers may have been placed on pharmacological or non-pharmacological treatments which may not yield significant benefit as their main concern hasn’t been addressed. Future research to test the assumptions in this theory are warranted.

## Conclusions

This novel substantive grounded theory of breastfeeding grief posits that the main concern of mothers with breastfeeding grief is unattained perception of motherhood, and that the core category, the process used to navigate this concern is reconciling motherhood ideology. Five interrelated stages of the core category including Responding to Disruptions, Disavowing Reality, Grieving, Accepting Reality, and Reframing Motherhood Ideology, were identified through which mothers move as they attempt to achieve emotional balance and reframe their motherhood ideology. Reconciling Motherhood Ideology is cyclical, not continuous, indicating how mothers re-experience previous emotions and reflections when a trigger is activated. The theory of breastfeeding grief provides a building block/framework for future studies examining breastfeeding grief and emphasizes that individuals can experience grief when they have experienced any form of loss (loss of human life or not).

## Data Availability

All data produced in the present study are available upon reasonable request to the author.

---

**Supplement 1.** Developing a Guideline for Reporting and Evaluating Grounded Theory Research Studies (GUREGT) checklist.

| Main area | Item | Question | Page number |
| --- | --- | --- | --- |
| Study aim | 1 | Is the grounded theory study aim presented to generate a theory of patterns of behaviour? | Yes. Introduction (Page 4) |
| Philosophical framework | 2 | Is the grounded theory embedded in any philosophical background? Why and how? | Yes. Methods > philosophical underpinning (Page 5) |
| The researchers' role | 3 | Is the researcher's theoretical sensitivity described according to conceptual thinking, level of insight into the research area and ability to generate meaning from data? | Yes. Methods > data analysis (Page 9) |
| Data collection | 4 | Is data collection methods described and explained? | Yes. Methods > data collection (Page 7-9) |
|  | 5 | Has qualitative or quantitative data collection methods been used? How and why? | Qualitative method was used because it is suitable for capturing sleep experiences from the perspective of participants using interview data.<br>Methods > design (Page 5-6) |
| Memos | 6 | Has memos been written throughout the study about concepts and categories and are they used to formulate and generate the theory? | Yes. Methods > data analysis (Page 7-8) |
| Sampling procedures | 7 | Is initial sampling conducted in the beginning of data collection described and explained? | Yes. Methods > participants (Page 5) |
|  | 8 | Is theoretical sampling of the emerging categories and theory from the data collection described and explained? | Yes. Methods > participants, inclusion and exclusion criteria, data analysis) (Page 6-8 and Supplementary material 2) |
|  | 9 | Is the selection of participants guided by theoretical sampling? How? | Yes. Methods > participants (Page 6-7) |
| Theoretical saturation | 10 | Is the reach of theoretical saturation explained according to no new insights relevant for the emergent theory? | Yes. Methods > inclusion and exclusion criteria (Page 6) |
| Analysis and coding | 11 | Is the coding levels and concurrent process of coding described according to open, selective and theoretical coding? | Yes. Methods > data analysis (Page 7-8) |
|  | 12 | Which concepts has guided the specific coding levels and how? | Yes. Methods > data analysis (Page 7-8) |
|  | 13 | Is the core category identified before conducting selective coding? | Yes. Methods > data analysis (Page 7-8) |
|  | 14 | Which theoretical codes have structured the theory to a progressive level of abstraction? | Yes. Methods > data analysis (Page 10) |
|  | 15 | Is the constant comparison method used to compare incidents with incidents, incidents with categories and categories with categories? | Yes. Methods > data analysis (Page 8-10) |
|  | 16 | Is the simultaneous data collection, analysis and coding guided by the theoretical sampling and writing memos described and explained? | Yes. Methods > data analysis (Page 8-10) |
| Review of literature | 17 | Is the literature reviewed avoided initially in the grounded theory study? Why and how? | Yes. Introduction (Page 4) |
|  | 18 | Is the literature reviewed after theory development on the basis of the emerging concepts and theory? How and on what grounds? | Yes. Introduction (Page 4) |
| Results/The theory | 19 | Is the main concern presented and explained? | Yes. Results > main concern (Page 12) |
|  | 20 | Is the core category and the related categories presented and explained? | Yes. Results > core category and factors accounting for variation in processing the main concern (Page 13-21) |
|  | 21 | Does the theory account for the overall pattern of behaviour in the substantive area? | Yes. Results > factors accounting for variation in processing the main concern<br>(Page 21 and Figure 1) |
|  | 22 | Is conceptualization used rather than description using quotes when writing the theory? | Yes. Conceptualization was used and some quotes were presented as indicators. |
|  |  |  | Results<br>(Page 13-21) |
| Discussion | 23 | Are the key relationships between the core category and concepts discussed and related to relevant literature? | Discussion<br>(Page 22-24) |
| Evaluation criteria | 24 | Are the criteria of fit, work, relevance, and modifiability presented and explained? | Yes. Methods > Rigor<br>(Page 10-11) |
|  | 25 | Are the evaluation criteria used to evaluate the theory? | Yes. Methods > Rigor<br>(Page 10-11) |

**Supplement 2.** Inclusion criteria modifications.

| **Purposive sample** | **Theoretical sample** |
| --- | --- |
| **Not included** | Race<br>AI/AN<br>Asian<br>Black or African/American<br>Native Hawaiian or other PI<br>White<br>Other, please specify_____ |
| **Not included** | Ethnicity<br>Hispanic, Not Hispanic, Other, please specify |
| Have you given birth to any children?<br>Yes, No | Do you have a biological child under 1 year?<br>Yes, No |
| Was there any time you planned to breastfeed your child(ren) but were not able to breastfeed at all?<br>Yes, No | Did you plan to breastfeed your child who is currently under 1 year?<br>Yes, No |
| Was there any time you planned to breastfeed your child(ren) but were not able to breastfeed at all?<br>Yes, No | Were you able to breastfeed your child who is currently under 1 year at all?<br>Yes, No |
| Was there any time you planned to breastfeed your child(ren) for a certain amount of time but couldn't breastfeed for as long as you wanted to?<br>Yes, No | For your child who is currently under 1 year, were you able to breastfeed as long as you wanted to?<br>Yes, No |
| Was there any time you planned to feed your child(ren) with breast milk only but had to introduce water, juice, formula or other foods earlier than you planned?<br>Yes, No | For your child who is currently under 1 year, did you plan to feed with breast milk only but had to introduce water, juice, formula or other foods earlier than you planned?<br>Yes, No |
| **Not included** | On a scale of 0-10, how personal is it to you that you were not able to meet your breastfeeding plan for your child who is currently under 1 year?<br>0 (Not personal)-10 (Extremely personal) |
| Are you experiencing/have you experienced any form of deep sadness or grief because you were not able to meet your breastfeeding plan(s) for your child(ren)?<br>Yes, No | Are you experiencing/have you experienced any form of deep sadness or grief because you were not able to meet your breastfeeding plan(s) for your child who is currently under 1 year?<br>Yes, No |
| Is the child(ren) because of whom you experienced breastfeeding sadness alive?<br>Yes, No | **Not included** |

**Supplement 3** Interview questions for theoretical sample

1) Can you please share with me your breastfeeding experience for your child under age 1?
2) Follow up: You mentioned you experienced (breastfeeding grief emotions), can you kindly walk me through how these occurred, like in stages? For those who didn’t mention any emotion: how do/did you feel about not meeting your breastfeeding goal?
3) Is any of these emotions related to how to see yourself as a mother?
4) You identified from the screening survey that not meeting your breastfeeding goals is/is not personal for you (score of 0-10), why did you choose the score?
5) What is your cultural practice regarding breastfeeding? Would you say your culture in any way contributes to how you are feeling/felt about not achieving your breastfeeding goal?
6) Can you tell me about your personality? Do you think your personality also affects how you are feeling/felt about not achieving your breastfeeding goal?
7) Is this your first child? If no, have you had previous unsuccessful breastfeeding experiences and you are attempting to ‘make it right’ with this child? If yes, do you think this contributes to how you are feeling/felt about not achieving your breastfeeding goal?
8) Were you really looking forward to breastfeeding? Can you share examples of what you did as you were preparing to breastfeed? Would you say your preparation/anticipation also contributes to how you are feeling/felt about not achieving your breastfeeding goal?
9) When you compare your experience with other moms who are able to mee their breastfeeding goals, or view breastfeeding contents online, does this make you react a certain way?
10) When you set your breastfeeding goal, were you very ‘strict’ with it or did you have alternative plans?
11) What support would you have loved when your emotions around not achieving your breastfeeding goal were very intense?
12) What aspect of your breastfeeding experience stood out for you that we have not yet talked about?

